# 1-C Nonlinear Covid-19 Epidemic Model and Application to the Epidemic Prediction in France

**DOI:** 10.1101/2020.05.24.20111807

**Authors:** J.-P. Quadrat

## Abstract

We have shown in a previous paper that the standard time-invariant SIR model was not effective to predict the 2019-20 coronavirus pandemic propagation. We have proposed a new model predicting *z* the logarithm of the number of detected-contaminated people. It follows a linear dynamical system *ż* = *b* − *az*. We show here that we can improve this prediction using a non linear model *ż* = *b* − *az^r^* where *r* is an exponent that we have also to estimate from data. Some countries have an epidemic with a bell shaped form that we call unimodal epidemic. With this new model, we fit observed data of different countries having an unimodal epidemic with a surprising quality. We discuss also the prediction quality obtained with these models at the epidemic start in France. Finally, we evaluate the containment impact on the Covid French mortality in hospitals.

## 1 Introduction

The forecast of the spread of the Covid epidemic is very important to take governmental decisions such as containment policy. In France, the decision of the population containment has been taken on the basis of possible risk of 500 000 deaths forecasted by Ferguson’s model. This model [4] is a renewal stochastic model using the SIR (Susceptible-Infected-Recovered) with a time-varying reproduction number that predicts the average number of secondary infections at a given time. In [2] we have shown that time-invariant SIR models were not effective to forecast the number of contaminated-detected people and we have proposed a new model. It was a dynamical linear model forecasting the logarithm of the cumulated infected-detected population *z*. The dynamics was *ż* = *b* − *az*. In this paper, we show that this model can be improved using the dynamics *ż* = *b* − *az^r^* where *r* is a real number to be estimated from observed data.

These models are good for what we call an unimodal epidemic which has a daily number of contaminated people with a bell shaped form. We will call these models 1-C, 1 for unimodal, and C for contaminated which correspond to the sum of I infected and *R* recovered of the SIR models. In [2] we have given an explicit formula for this form that is 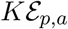 where 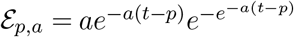 is a probability density without moments but with a peak at *p* and *a* controlling the width of the bell shape. Probably, the general form of an epidemic can be described as a linear finite or infinite mixture of such densities that we call epidemic probabilities. Here, we introduce this *r* number which controls the right probability queue of the bell shape. But we are not anymore able to give an explicit form of the corresponding probability density.

Based on this model able to predict the infected people number, and the fatality rate observed in France, we can compute an estimator of Covid deaths in hospitals that would have happened without the containment policy and compare it with the prediction given by [5]. But, to improve the confidence in the obtained figures, we have to discuss about the robustness of the predictor at the epidemic start. Indeed, at the beginning the noise on data is large and a small error on the estimated parameters *a, b* and *r* can produce a large error on the size of contaminated people at the end of the epidemic.

## 2 The nonlinear model

The nonlinear model has been derived from the criticism of the standard SIR model which, in its time-invariant version, was not able to follow the observed data. Let us recall briefly the criticism done in [2] to be self-contained.

The SIR (Susceptible-Infected-Recovered) model distinguishes three kinds of people in a given population. It denotes by *S* the proportion of the susceptible population to be infected, *I* the proportion able to infect and by *R* the proportion which has recovered and which is not able to transmit the infection. We have *S + I + R* = 1. This model assumes that the number of newly infected people is proportional to the product of the number of infected by the number of susceptible ones. Based on this assumption, the following differential equations describing the evolution of the three variables are proposed:

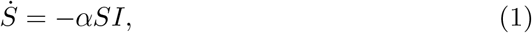

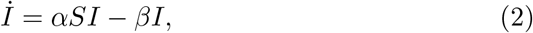

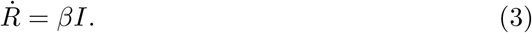

Where *α* can be seen as a reproduction number by a unit of time and *β* is a recovering factor by a unit of time. From a given epidemic, to forecast its evolution, we have to estimate the two numbers *α* and *β*.

Using the new variable *C* = *I* + *R*, instead of *I*, then with *S* = 1 − *C* and *I* = *C − R* the system can be written as:

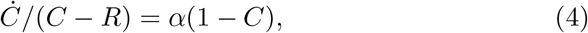

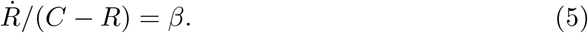

This form is useful to estimate parameters *α* and *β*. The variable *C* will be named *contaminated people*.

For a given epidemic, to be able to forecast the evolution, we have to estimate the two numbers *α* and *β*. The problem is that we are not able to observe any of the two variables *C, R*. We can test some people or observe some symptoms but we cannot observe precisely the number of contaminated people.

At the beginning of the infection, as nobody has recovered, we can suppose that *R* = 0 (which corresponds to *β* = 0). If we suppose that the number of contaminated-detected people is proportional to the contaminated ones (*y* = *NC* where *N* is an unknown parameter and *y* denotes the contaminated-detected cases) we obtain the evolution equation of *y*:

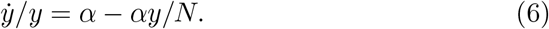

This equation can be written:

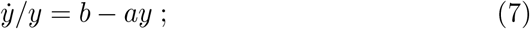

with *b* = *α* and *a* = *α/N* which can be easily estimated from the observation *y* by a simple linear regression.

Let us see the results on French data for the covid epidemic. The Figure-1 gives the observation in green of *ẏ/y* and in black its regression *b − ay*(*t*). We cleary see the obtained result is poor. A better regression is obtained by the blue line with *B − A*log(*y*(*t*)).

**Figure 1:**
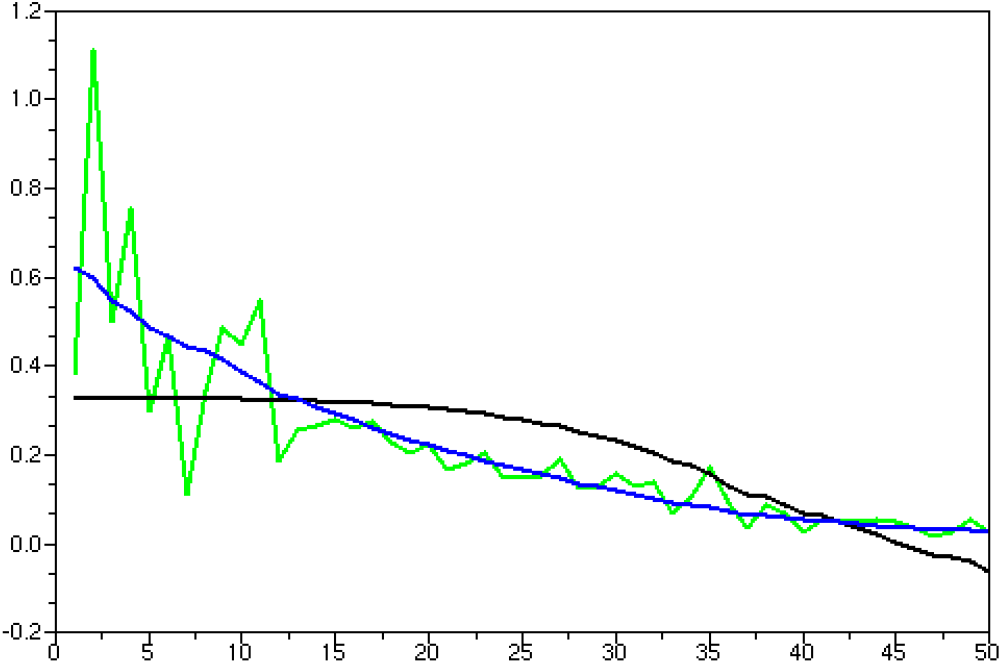
Green: observed *ẏ*(*t*)/*y*(*t*). Black: *b−ay*(*t*). Blue: *B − A* log(*y*(*t*)).

Therefore, a better model than the SIR model is given by:

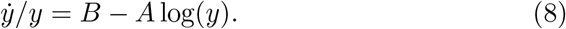

If we see the term “−*A* log(*y*)” as a brake on the exponential epidemic growing, it cannot be simply explained in immunology terms as the standard SIR brake “−*ay*”. While it is not clear to understand its origin, it probably comes from a more local notion of susceptible-to-be-infected people to be defined. With particle-based simulation methods [3] it is perhaps possible to understand this term.

Using the new variable *z* = log(*y*) we obtain the simple model *ż* = *B* − *Az*. In [1] Bhardway has proposed a linear dynamical model: *ẏ* = *e^−at+b^y*, which gave the same final parameterization.

Based on the availability of more and more data with time it appears that we can improve the fit using the following model:

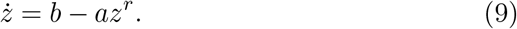

Let us give the results obtained, in the Germany case, for different values of *r*: Figure-2a *r* = 3, Figure-2b *r* = 2 Figure-3a *r* = 1 and also for the model SIR *ż* = *b − ae^z^* Figure-3b.

**Figure 2:**
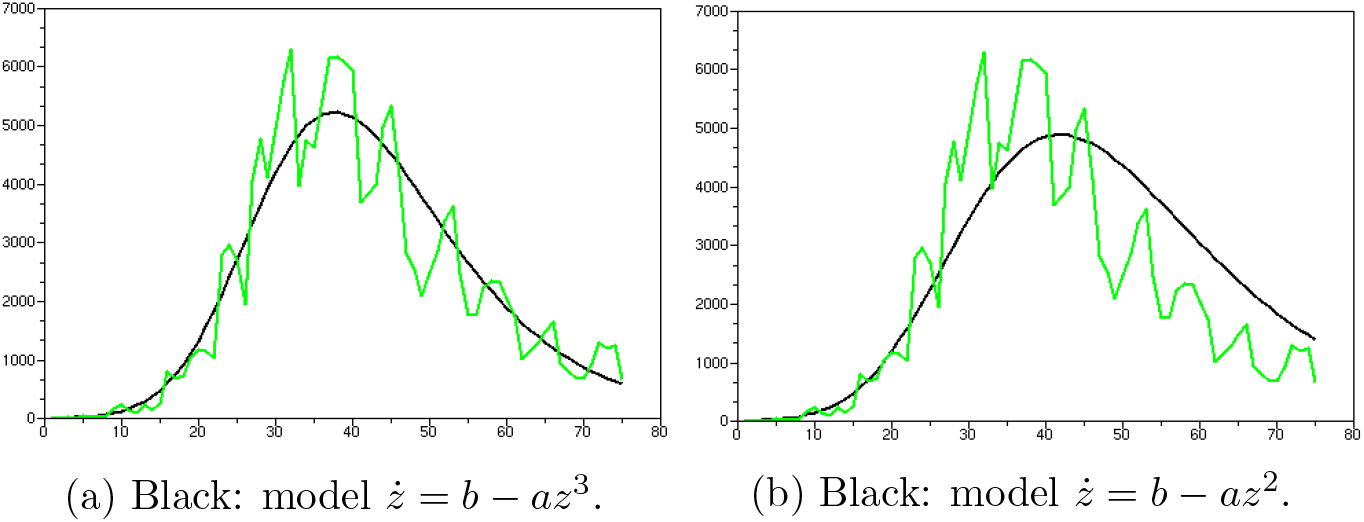
Daily Contaminated Germany in green.

**Figure 3:**
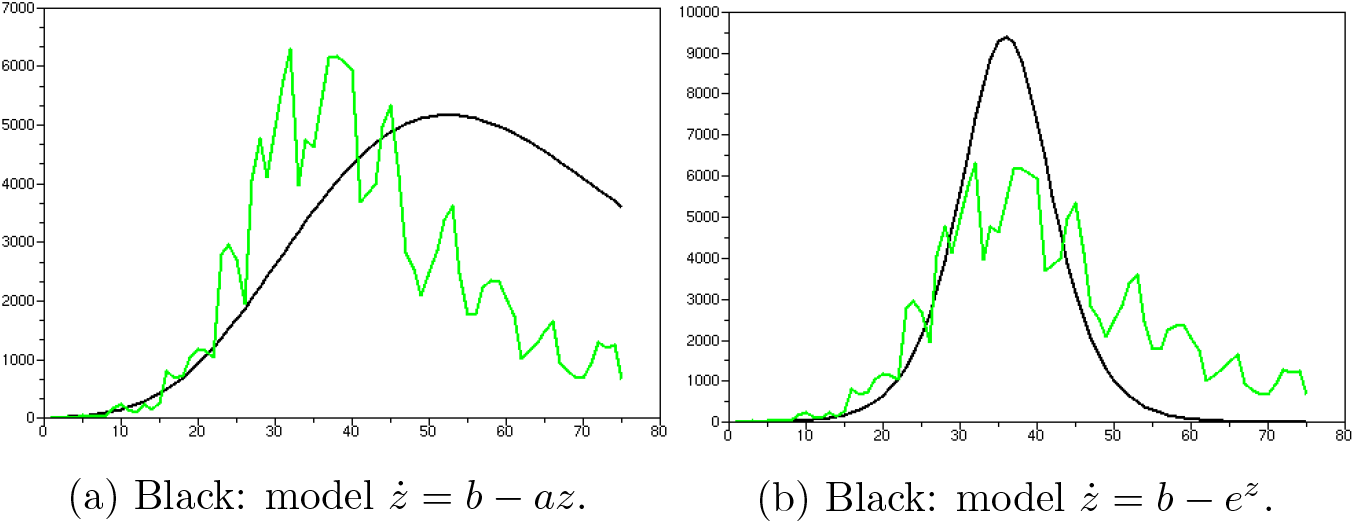
Daily Green: Daily contaminated people in Germany.

The linear model *r* = 1 is good only locally in time. In [2] we gave the result obtained with the linear model based on the 30 last observed data. The fit of the linear model was as good as it is now at the end of Figure-2a with *r* = 3. We see here that the fit gives greater importance to the first data. It is not surprising because the noise at the beginning is much larger than at the end of the epidemic and therefore its contribution to the regression error is larger.

It is quite surprising that we can fit completely an epidemic with only three parameters *a, b, r* and its starting time. In the future, we will try to understand the origin of this exponent because its meaning is currently unclear.

Therefore we have:

#### THE MAIN POINT OF THE PAPER

The logarithm of the observed number of people contaminated during an unimodal Covid epidemic denoted by *z*(*t*) can be well approximated by a first-order time-invariant stable nonlinear dynamical system:

*ż* = *b* − *az^r^* with *a >* 0.

Let us give also the results obtained for France in Figure-4a (*r* = 2) and Italy in Figure-4b (*r* = 1.7).

**Figure 4:**
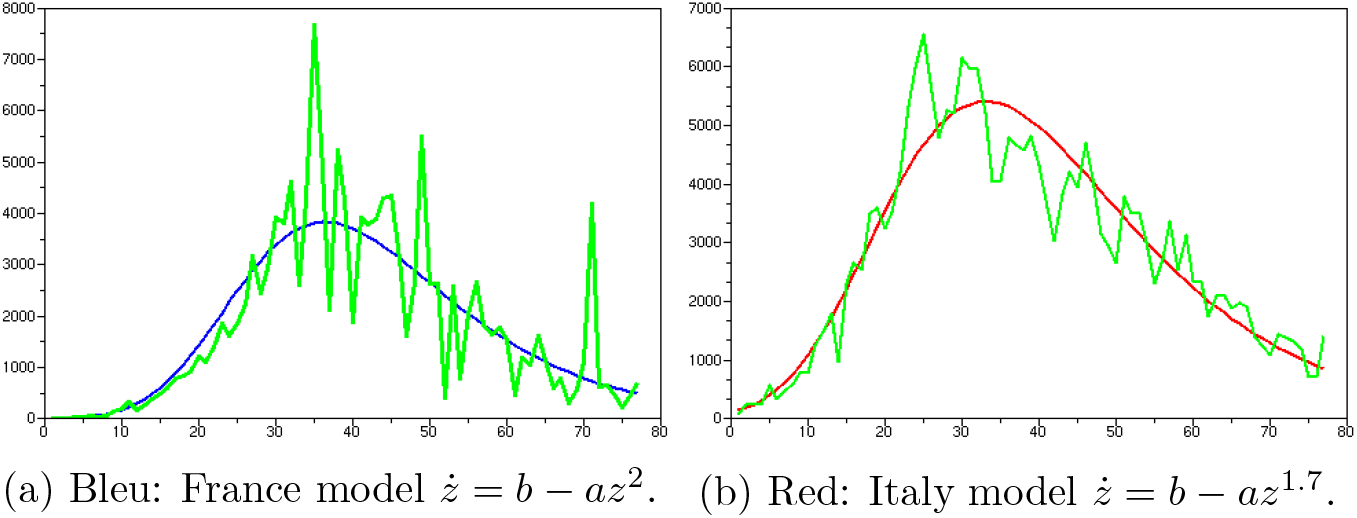
Green: Observed daily contaminated people in France and Italy (last observation made in May 12, 2020).

It is easy to estimate the parameter *r* at the end of the epidemic. To take the right political decision it is important to have a good prediction at the beginning. Let us see in the next section what we can obtain with such models.

## 3 On the prediction robustness

The role of models is their abilities to provide a prediction which can help to take political decision such as containment. In [2] we have used the linear France model (*r* = 1) to see the impact of the French containment. It seemed that our prediction based on data before containment was reliable because it was consistent with the results based on data after containment. It appears later than the prediction before containment was quite sensitive to the numbers of taken data. Our prediction of 250 000 contaminated people detected in hospital can increase to 400 000 by adding four or five data. This last prediction seems to us more reliable. It does not change the conclusion of the paper [2], but we want to revisit the prediction of the contaminated people detected in hospitals, at containment decision date, trying to be more robust, knowing that the nonlinear model is a much better model than the linear one.

The main difficulty is the size of the noise which appears at the start of the epidemic. The newly detected people are much larger in proportion at the beginning than at the end (see Figure-1). With such a level of noise, the difference between the linear (*r* = 1) and the quadratic case (*r* = 2) is not significant if we restrict the set of data to the ones available before containment (see Figure-5a). To improve the quality of the regression we can filter the data before carrying out the regression. See the results on Figure-5b. The filtering has consisted in averaging on 7 days. We see, once more, that the two cases *r* = 2 and *r* = 1 do not make any significant difference. We can conclude that at the start of the epidemic we are not able to estimate the parameter *r*.

**Figure 5:**
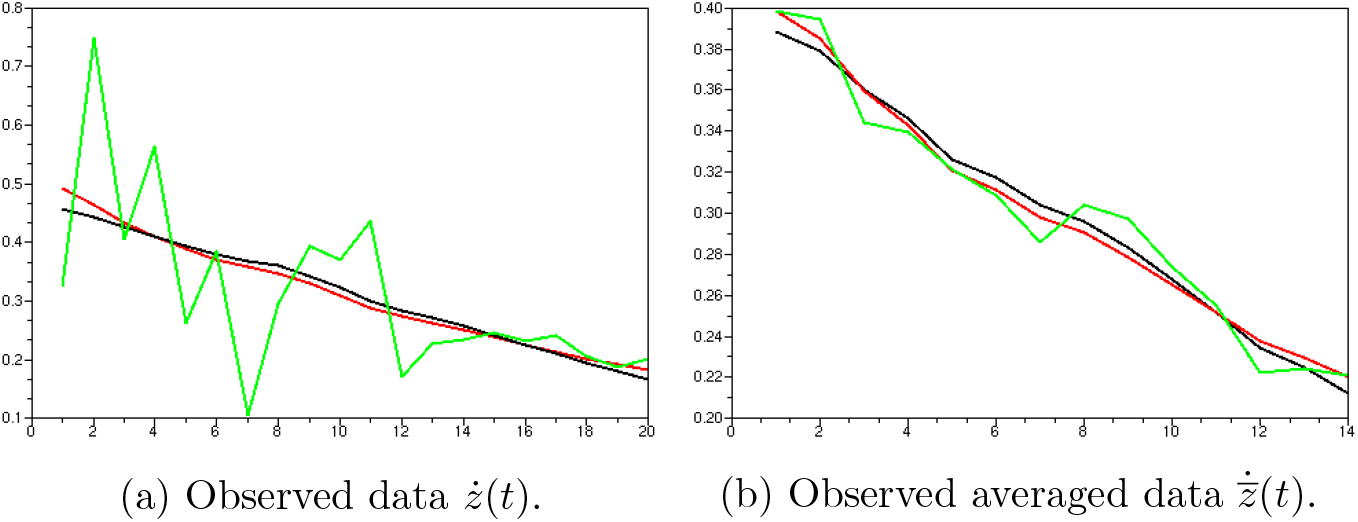
Regression on pre-containment data, black: *b − az*^2^(*t*), red: *b − az*(*t*).

Now, let us discuss a little bit about the robustness of the predictors. For that, we consider the prediction, obtained in the French case, of the cumulated infected people at the end of the epidemic in different cases. These quantities can be easily obtained. Indeed all the models considered here have an asymptote when t goes to infinity. It is given by 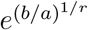 for the new proposed model and *b/a* for the SIR model. We obtain the following results:

1. 195 000 for pre-containment data with model *r* = 1,
2. 44 000 for pre-containment data with model *r* = 2,
3. 465 000 for pre-containment data with model *r* = 1 on weekly averaged data,
4. 67 000 for pre-containment data with model *r* = 2 on weekly averaged data,
5. 9 100 for pre-containment data with SIR model,
6. 144 000 for all data before May 13 with model *r* = 2,
7. 141 000 for the observed infected people in hospitals on May 13.

Firstly we have no confidence in the prediction of the SIR model. We have seen in Germany’s case that it was not good. We see here that its prediction is very small. We can see on the Web some good predictions based on SIR model but they are time variant SIR model. It is clear that if we adapt the SIR model in time it will make a reasonable short term prediction.

Secondly the nonlinear models give too small quantities. In fact the good *r* is obtained by data at the end of the epidemic because the *r* controls well the speed of convergence towards the asymptote. At the beginning of the epidemic it does not make a big difference and perhaps is worse than the linear model. Moreover a small difference in *r* can produce a large difference on the asymptote 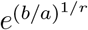.

Therefore we have only to discuss the cases 1 and 3, the linear model with data filtered or not. Since we have few data it seems more reliable to choose the case 3. It is well known in robust statistics that it is better to use estimator eliminating extreme data even if this elimination can reduce the performance of the estimator. Therefore for the sake of robustness we choose the linear model based on filtered data.

At the decision time we have an observed fatality rate of 0.02 see Figure-7 at day 20 (coherent with Japan fatality rate, the Chinese one was 0.04, etc…). Therefore the case 3, in the absence of containment policy, predicts 10 000 deaths (like the standard influenza). French government decided containment based on the Ferguson’s model predicting a risk of 500 000 deaths (50 times larger than our prediction).

## 4 Containment decision and its a posteriori evaluation

Prediction 1 is closer to the observed number of contaminated at present time (case 7) which is closed to the prediction 6 obtained with the complete set of data (case 6). But we think that the most reliable pre-containment prevision was the third one (*r* = 1 on filtered data) for two reasons.

- The first reason is that before the containment decision all the contaminated people are searched actively. After containment, only severe cases requiring hospitalization are detected. Severe cases represent only roughly 20 percent of the positive cases. Therefore a prediction of 465 000 positive cases corresponds well to 140 000 severe cases.
- The second reason comes from the following Figure-6 which shows the asymptote level as a function of time-based on a moving regression on a set of 40 observations filtered by averaging on 14 days. We see a regular reduction of the prediction of the number of contaminated people which agrees on the fact that the severe case part increase in the detected people. If we try to do the same thing on the unfiltered data, the curve would be a little bit chaotic. Indeed a bad data can have a huge influence on the predicted asymptote.

**Figure 6:**
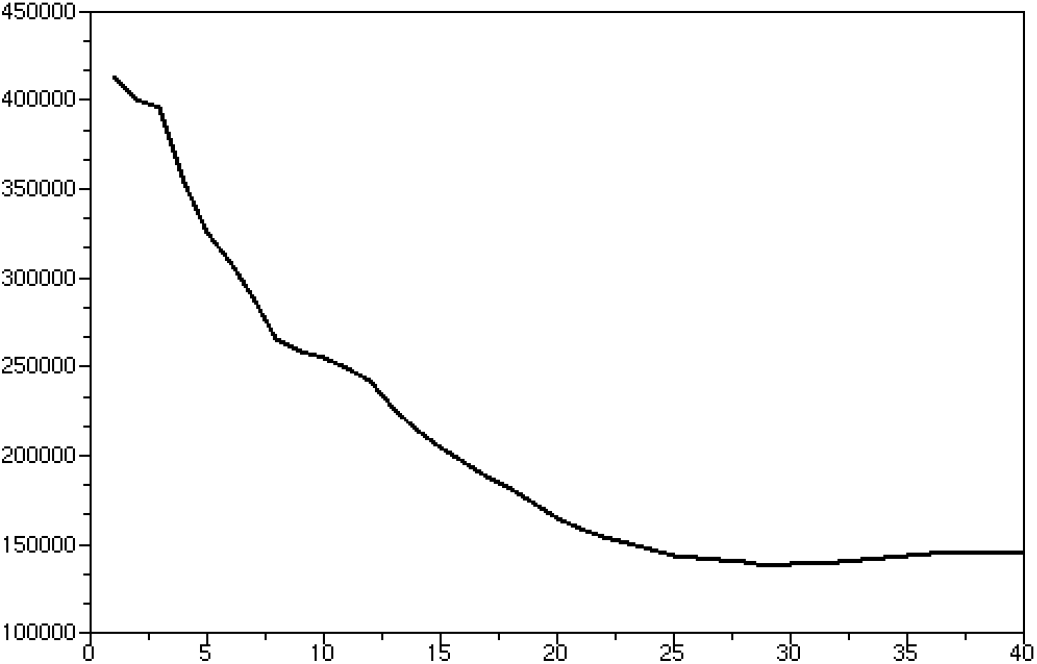
Predicted infected people as time function based on a moving regression.

The prediction of the number of contaminated people avoided is not 465 000 − 145 000 = 320 000 because the 145 000 are severly contaminated cases and 465 000 are contaminated-detected cases with an active test policy. If we think that the severe cases represent 20 percent of the detected cases, we see that the population containment seems to have increased the contaminated people. This surprising fact: that containment does not necessarily reduce the number of contaminated people, is confirmed by the preliminary study [10] where it is shown that workers contained have been more contaminated than the non contained ones.

Based on this more argued prediction than the one given in [2] and the French fatality rate as a time function given in Figure-7 we can estimate the number of deaths in the absence of containment. We have seen on day 21 on this figure that the fatality rate is approximately equal to 0.02 which gives 10 000 deaths in hospitals. This result has to be compared with the 60 000 death avoided predicted in [5] based on sophisticated SIR models. The observed death number in hospitals is 17 000 at the date of May 13. This estimation seems to be optimistic but it depends on the fatality rate used which needs to be discussed.

**Figure 7:**
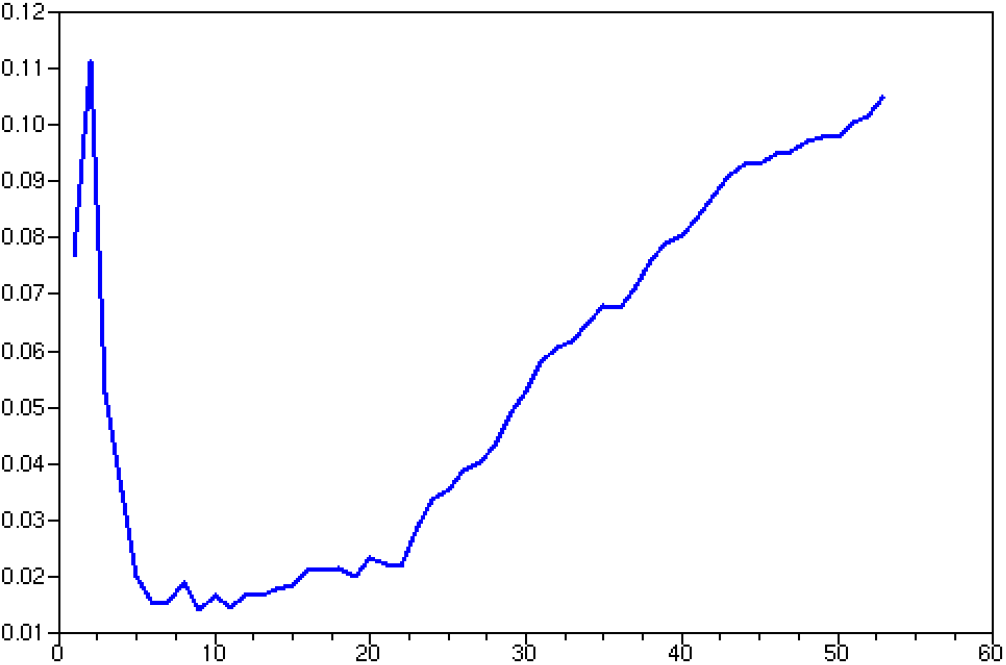
Covid fatality rate in France as a time function.

At present time the fatality rate in France (0.15) is the largest in the world. If we compute it with death and contaminated counted in the hospitals it becomes 0.12. The reason of this high fatality rate comes from the decision taken in phase^1^ 3, after containment decision, to treat only people with intensive respiratory distress requiring hospitalisation despite Pr. Raoult’s indications proposing an early treatment at this date [9]. Now Pr. Raoult has shown that with his treatment we can obtain a fatality rate of 0.005 for a population actively detected (that was the case for all the French population in phase 2). Therefore this prediction of 10 000 deaths without containment policy is probably over estimated by a factor 2 or 3 of what would have happened in absence of containment but with an active detection and an early treatment policy as the one proposed by Pr. Raoult. This abrupt conclusion must be moderated by two points: – firstly the screening tests were not available in France to apply Pr. Raoult’s policy easily, – secondly the prediction done here is based on an unimodal model, but without containment policy there is a risk, probably avoidable by the Pr Raoult’s policy, that this kind of unimodal model is not anymore valid (in the Sweden’s case a plateau is appearing at the top of the bell shaped form of the daily detected people).

## 5 Conclusion

SIR models are not effective to estimate the Covid epidemic. An alternative way to obtain a correct prediction is to fit a nonlinear time-invariant firstorder dynamical model *ż* = *b* − *az^r^* on the logarithm of the number of observed contaminated people. Moreover, we have shown that it is difficult to estimate *r* at the early stage of the epidemic and furthermore that it can be a bad idea to try to estimate it. A better idea was to fit a linear (*r* = 1) model on filtered data. Doing that, we obtained a more robust predictor. Once more time we conclude to, at least for the French’s case, that the containment was a bad idea and that a better idea would have been to follow the Pr. Raoult advice: test and treat without confining people.

## Data Availability

Wikipedia

## Footnotes

1 The three phases of the French Covid policy were: – in phase 1: search the first positive case, – in phase 2: no longer try to find anymore the initial case but all the positive ones, – in phase 3: no longer try to find all the positive cases but only the severe ones that require hospitalization.

